# Precision prescribing of SGLT2-inhibitors in people with type 2 diabetes for primary prevention of heart failure: model development and validation study

**DOI:** 10.1101/2025.09.08.25335314

**Authors:** Katherine G Young, Andrew P McGovern, Rhian Hopkins, Thijs T Jansz, Pedro M Cardoso, Rury R Holman, Ewan R Pearson, Andrew T Hattersley, Angus G Jones, Kieran Docherty, Naveed Sattar, Beverley M Shields, John M Dennis, the MASTERMIND Consortium

## Abstract

**Objectives:** To develop and validate a model to predict individual-level benefits of SGLT2-inhibitors for heart failure prevention in people with type 2 diabetes who do not have atherosclerotic cardiovascular disease (ASCVD), heart failure (HF) or chronic kidney disease (CKD).

**Design:** Population based cohort study.

**Setting:** The SGLT2-inhibitor absolute response (SABRE) model estimated individual-level 5-year heart failure benefits using absolute heart failure risk predictions from the previously validated QDiabetes-Heart Failure model, combined with the hazard ratio of SGLT2i-associated hospitalisation for heart failure benefit (HR: 0.63 [95% CI: 0.50 to 0.80]) from trial meta-analysis. The data source was primary care records with linked hospital admission and death records (Clinical Practice Research Datalink, England) from January 2013 to October 2020.

**Population:** Adults with T2D without ASCVD, HF or CKD initiating an SGLT2i (n=57,368) or, as a comparator group, DPP4-inhibitors or sulfonylureas (n=111,673). Propensity score weighting and regression adjustment were used to mitigate potential treatment selection bias. Validation of model accuracy was performed by comparing the calibration of predicted versus observed heart failure benefits in those receiving SGLT2i versus comparator therapy.

**Main outcome measures:** New-onset heart failure recorded in primary care, linked hospital or death records.

**Results:** Amongst 57,368 SGLT2i and 111,673 DPP4i/SU treatment initiations, estimated risk of new-onset heart failure was 30% lower with SGLT2i versus the comparator arm (HR: 0.70 [95% CI: 0.63 to 0.78]), similar to trial meta-analysis for hospitalisation for heart failure. The relative benefit of SGLT2i on heart failure was consistent across all levels of baseline absolute heart failure risk as estimated from the QDiabetes-Heart Failure model (p=0.82 for treatment arm:baseline heart failure risk interaction). 5-year absolute heart failure benefit predictions with SGLT2i ranged from <0.1% to 14.1% across individuals (median 1.0% [IQR 0.6-1.8%]), and predictions were well-calibrated against observed heart failure outcomes. Model evaluation established that individualised targeting of SGLT2i initiation using SABRE could more efficiently reduce heart failure outcomes compared to current treatment recommendations for those with T2D without ASCVD/HF/CKD.

**Conclusions:** SGLT2i can be targeted to individuals with type 2 diabetes without ASCVD/HF/CKD for primary prevention of heart failure using an easily deployed clinical prediction model integrating evidence from clinical trials.

## INTRODUCTION

SGLT2-inhibitors (SGLT2i) substantially reduce the risk of hospitalisation for heart failure and, to a lesser extent, reduce atherosclerotic cardiovascular disease (ASCVD) events in people with type 2 diabetes (T2D) ^1^. Guidelines consistently recommend SGLT2i for individuals with T2D who have established ASCVD or history of heart failure (HF) ^2^ ^3^, who have the highest absolute risk of both ASCVD events and heart failure and are therefore likely to derive the greatest benefit. However, in those with T2D without ASCVD or HF, evidence from trial meta-analysis of SGLT2i-associated ASCVD benefit is uncertain ^1^. This group represents around two-thirds of those with T2D in the UK ^4^.

For those with T2D without ASCVD/HF, international (ADA/EASD) guidance suggests the use of SGLT2i in those aged ≥55 years with the presence of 2 or more established cardiovascular risk factors including obesity, hypertension, or dyslipidaemia ^2^. In contrast, UK guidance for SGLT2i targeting in this group is now based on an individual-level predicted 10-year ASCVD risk of ≥10% ^3^, a threshold exceeded by 89.6% of people with T2D without ASCVD/HF in the UK on anti-hyperglycaemic treatment ^4^. Beyond these broad recommendations, current guidance does not give advice regarding which people with T2D without ASCVD/HF are likely to have the greatest cardiovascular benefit if treated with SGLT2i.

We aimed to develop and validate an ‘SGLT2i absolute benefit response’ model (SABRE) to predict the absolute benefit of SGLT2i for primary prevention of heart failure for individual patients with T2D without ASCVD/HF. We followed the risk-modelling approach outlined in the PATH (Predictive Approaches to Treatment effect Heterogeneity) statement ^5^. Individual-level predictions were derived by combining the RCT-estimated relative risk reduction of SGLT2i for heart failure prevention, and absolute heart failure risk estimates from an established validated clinical prediction model ^6^. Accuracy and clinical utility were assessed in a large population-representative UK dataset of people with T2D.

## METHODS

### Study population

The study population was identified using longitudinal UK primary care data from Clinical Practice Research Datalink (CPRD) Aurum ^7^, including those with valid linkage to national hospital inpatient data (Hospital Episode Statistics), Office for National Statistics (ONS) death data and patient Index of Multiple Deprivation (IMD) data. CPRD covers around 20% of the UK population and is broadly population-representative ^7^ ^8^.

The study population was those with type 2 diabetes (T2D), with no atherosclerotic cardiovascular disease (ASCVD), no history (ever) of heart failure, and no chronic kidney disease (CKD; stage 3-5) or macroalbuminuria in primary care or hospital records (i.e. excluding those for whom evidence-based guidance for SGLT2i treatment already exists). We included those initiating SGLT2i, or as a comparator, those initiating DPP4-inhibitors (DPP4i) or sulfonylureas (SU), excluding those already receiving SGLT2i, between 1 January 2013 and 31 October 2020. Treatment initiation was defined as the first-ever prescription for each drug class. The date of first prescription defined the study baseline and time zero for follow-up. We excluded people receiving a GLP1-receptor agonist or thiazolidinedione at baseline as these treatments may modify heart failure risk ^9^ ^10^. We also excluded people aged<25 or >84 years at baseline as both groups were outside the range of the prediction model for new-onset heart failure. We further excluded individuals prescribed SGLT2i, DPP4i or SU as first-line treatment as this is not recommended in UK guidelines ^3^, and drug initiations in the 90 days following registration at the GP practice as these may represent continuation of existing treatment where a patient has transferred from a different GP practice. Codelists and implementation algorithms for defining the study population and baseline characteristics are described here: https://github.com/Exeter-Diabetes/CPRD-Codelists/tree/pre-2024 (additional detail in ESM Methods).

### Treatment comparison

DPP4i and SU (pooled) were used as comparator drug classes as there is little evidence that either impact heart failure risk beyond their glucose-lowering effects ^11–13^ (the DPP4i saxagliptin and the older SUs chlorpropamide and tolbutamide may increase heart failure/CVD risk ^14^ ^15^ but these accounted for <4% of comparator group initiations in our study). We confirmed the validity of pooling both drug classes in our study population (see Sensitivity analyses).

Individuals were followed from treatment initiation to the end of the follow-up period, defined as the earliest of: the date of death, deregistration from primary care, the last collection date from their GP practice (maximum 15/10/2020), or five years post-drug initiation. We censored individuals if they initiated GLP1-receptor agonist or thiazolidinedione therapies due to their impact on heart failure risk. To ensure no overlap in follow-up time between comparator treatment arms, individuals in the SGLT2i arm were censored if initiating DPP4i or SU, and individuals in the DPP4i/SU arm were censored if initiating SGLT2i. Individuals contributed follow-up time regardless of whether they adhered to the treatment after initiation. Alternative censoring strategies were tested in sensitivity analyses.

### Outcome

The primary outcome was new-onset heart failure, defined as the earliest code for heart failure in primary care, hospital records or mortality records. This is a broader definition than the trial endpoint of hospitalisation for heart failure only, which we assessed in sensitivity analysis.

### Heart failure and CVD risk scores

5-year absolute heart failure risk predictions were derived from QDiabetes-Heart Failure (QDiabetes-HF) 2015 ^16^. QDiabetes-HF was developed and validated in UK primary care data for the same primary outcome as above on over 500,000 patients with type 1 or 2 diabetes from 1998 to 2014, prior to any widespread use of SGLT2i. The model includes age, sex, ethnicity (9 groups derived from the 16 UK census categories: White, Indian, Pakistani, Bangladeshi, Black Caribbean, Black African, Chinese, Other), deprivation score, smoking status, diabetes type, diabetes duration, history of heart attack/angina/stroke, atrial fibrillation, chronic kidney disease stage 4 or 5, HbA1c, total cholesterol:HDL ratio, systolic blood pressure (SBP) and BMI as predictors. Cholesterol:HDL, SBP and BMI were imputed from age, sex, ethnicity, and smoking status where missing. We also derived the predicted 10-year cardiovascular risk from QRISK2 2017 ^17^, a validated UK general population prediction model for new-onset cardiovascular disease (myocardial infarction, stroke, angina, transient ischaemic attack). Detailed variable definitions can be found in ESM Methods.

### Weighting

To control for potential treatment selection bias, overlap weighting using propensity scores ^18^ and regression model adjustment were used. The full covariate set used for both propensity scores and adjustment included: age, sex, ethnicity, deprivation score, smoking status, diabetes duration, atrial fibrillation, HbA1c, SBP, BMI, absolute heart failure risk (from QDiabetes-HF), hypertension, number of emergency inpatient hospital admissions in the previous year, number of glucose-lowering drug classes ever prescribed, number of other current non-insulin glucose-lowering medications, current insulin use, and year of current drug (SGLT2i/DPP4i/SU) initiation (see ESM Methods for detailed variable definitions). For regression model adjustment, continuous covariates were modelled as 5-knot cubic splines to allow for non-linearity. All analyses were complete case due to the limited number of individuals with missing data (ESM Figure 1).

**Figure 1:**
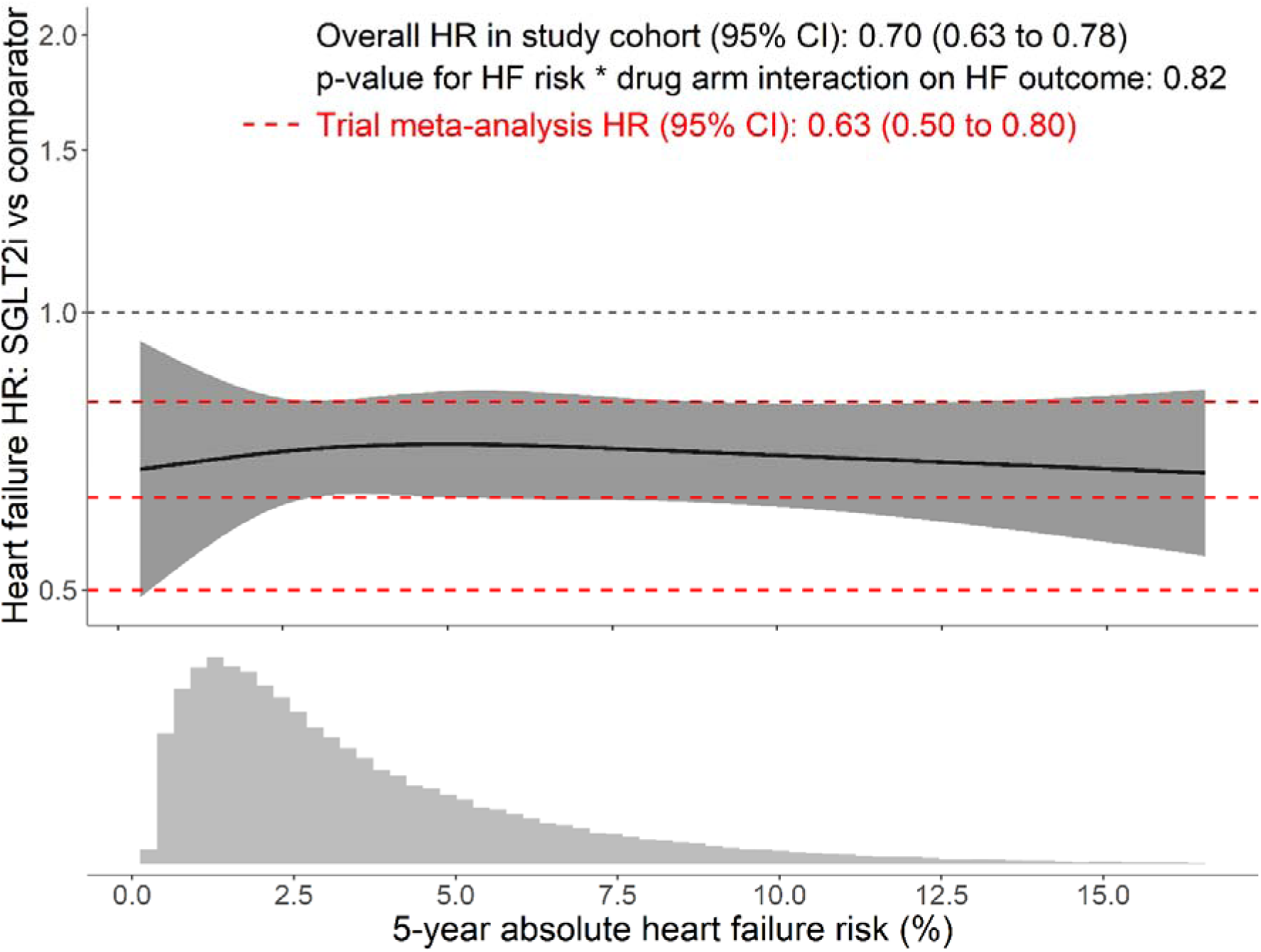
Hazard ratio estimates for new-onset heart failure for SGLT2-inhibitors vs comparator (DPP4-inhibitors/sulfonylurea) across baseline heart failure risk (model developed on n=169,041 [whole study cohort] using QDiabetes-Heart Failure for 5-year absolute heart failure risk estimates). Solid black line represents estimated hazard ratio (HR) at a given level of 5-year heart failure risk; grey shading represents 95% CI. Red dashed lines represent trial meta-analysis result with 95% CI. ‘Drug arm’ = SGLT2-inhibitor or comparator (DPP4-inhibitor/sulfonylurea). Lower plot shows distribution of baseline 5-year heart failure risk estimates within the study cohort. HF=heart failure, HR=hazard ratio, SGLT2i=SGLT2-inhibitor

### Statistical analysis

#### Evaluation of accuracy of SABRE model inputs

We assessed the calibration of absolute 5-year heart failure risk predictions from QDiabetes-HF by comparing predictions to the observed incidence of heart failure in the comparator (DPP4i/SU) arm only, stratified by decile of predicted risk. Observed incidence was estimated by fitting an unadjusted Cox proportional hazards model with risk score decile as the only predictor to all individuals, and using this model to estimate heart failure-free survival at 5 years for each decile.

A meta-analysis of the 3 major SGLT2i trials in those with T2D and no ASCVD or HF (>90% without history of HF ^19–21^) has shown a relative risk reduction for hospitalisation for heart failure of 37% (HR: 0.63 [95% CI: 0.50 to 0.80]) ^1^. This meta-analysis did include up to 25% of patients^19^ ^22–25^ with CKD and/or macroalbuminuria, unlike our study population where this group was excluded as evidence-based SGLT2i treatment guidance already exists for these groups. To assess whether the benefit of SGLT2i from this meta-analysis was the same for a broader heart failure outcome in our study population, we estimated the overall hazard ratio using adjusted Cox proportional hazards models with overlap weighting based on propensity scores (see ‘Covariates’ for variables used).

We also explored whether the hazard ratio for heart failure benefit with SGLT2i varied by baseline absolute heart failure risk. Non-linear continuous hazard ratios were estimated using an adjusted Cox model incorporating a drug arm (SGLT2i or comparator) by risk score (modelled as a 3-knot restricted cubic spline) interaction term.

#### Estimation and validation of absolute SGLT2i heart failure benefits from SABRE

To derive individual-level predictions of 5-year absolute heart failure benefit with SGLT2i, individual 5-year heart failure risk predictions from QDiabetes-HF were multiplied by the relative SGLT2i risk reduction on heart failure from trial meta-analysis (37%). For example, for a patient with a 5-year QDiabetes-HF score of 20%, SGLT2i are predicted to reduce this risk by 37%, equating to an absolute benefit of 7.4% (37% of 20%).

Validation of predicted benefits at the individual patient level was not possible as the counterfactual outcome (the outcome an individual would have had if they had received the alternative treatment) was unobserved. As an alternative, we estimated individual-level observed benefits by fitting an overlap-weighted and adjusted Cox model (using the full covariate set, see ‘Covariates’) to the entire cohort, incorporating a term for treatment (SGLT2i versus comparator), and used this to predict counterfactual observed outcomes on both SGLT2i and comparator for each individual. Observed benefit was then estimated for each individual as the difference in predicted 5-year heart failure risk between SGLT2i and comparator. To compare predicted and observed benefits, individuals were grouped by decile of predicted benefit from SABRE and validation performed by comparing median predicted and median observed benefits of individuals within each decile.

#### Evaluation of 7 treatment strategies based on current treatment guidelines and SABRE predictions

7 potential binary (treat SGLT2i, do not treat SGLT2i) treatment strategies based on current guidelines and the SABRE model were defined as follows:

1: UK NICE guidance: treat individuals with QRISK2 score >10% ^3^.

2: International ADA/EASD guidance: treat individuals aged ≥55 years with 2 or more risk factors, including obesity, hypertension, or dyslipidaemia ^2^.

3: SABRE model matched to NICE: treat individuals above a threshold of predicted heart failure benefit that results in recommending SGLT2i to the same proportion of individuals as NICE guidance.

4: SABRE model matched to ADA/EASD: treat individuals above a threshold of predicted heart failure benefit that results in recommending SGLT2i to the same proportion of individuals as ADA/EASD guidance.

5: SABRE model restricted strategy: treat individuals above a more restricted threshold than strategy 4, corresponding to the top 25% centile of predicted heart failure benefit in those recommended SGLT2i by ADA/EASD guidance.

6: QRISK2 matched to ADA/EASD: treat individuals above a QRISK2 threshold that results in recommending SGLT2i to the same proportion of individuals as ADA/EASD guidance.

7: QRISK2 restricted strategy: As QRISK2 is the widely used CVD risk score in UK clinical practice ^26^, repeat strategy 5 but using a QRISK2 threshold (instead of SABRE) which results in recommending SGLT2i to the same proportion of individuals as strategy 5.

We then calculated expected outcomes for each treatment strategy (alongside a ‘treat all’ strategy), comprising the proportion of the cohort who were SGLT2i treated, the absolute benefit in those treated (number of HF events prevented per 100 patient-years), and the number needed-to-treat per year to prevent one HF event (NNT). 95% confidence intervals for absolute benefits were estimated using bootstrap resampling (1000 iterations). For each strategy, we also estimated observed SGLT2i benefits as the difference in overlap-weighted 5-year cumulative incidence of heart failure with SGLT2i versus the comparator arm, fitted separately for those indicated and not indicated for treatment.

#### Sensitivity analyses

Sensitivity analyses were performed to test the robustness of our estimate of the hazard ratio for heart failure benefit with SGLT2i, as follows:

1. Assessment of the validity of the pooled comparator arm by evaluating heart failure outcomes with SGLT2i versus SU-only and DPP4i-only comparator arms.
2. Assessment of the method used to adjust for potential treatment bias, by testing: a) use of overlap weighting only without multivariable adjustment, b) multivariable adjustment without weighting, and c) inverse probability weighting with multivariable adjustment.
3. Modification of the primary outcome definition, by repeating the SGLT2i-comparator arm comparison using a more specific definition of hospitalisation or death with heart failure as the primary cause, which more closely resembles the outcome used in clinical trials.
4. Inclusion of only individuals initiating second-line therapy after metformin (so individuals could only be in one of the SGLT2i, DPP4i or SU groups), with a) an intention-to-treat approach (no censoring due to glucose-lowering treatment change), and b) a per-protocol approach (censoring at earliest glucose-lowering treatment change including stopping current drug).
5. Treatment of death from non-heart failure causes as a competing risk, to investigate the possible impact of informative censoring.
6. Inclusion of only individuals co-treated with insulin to investigate possible effects of insulin treatment on SGLT2i-associated heart failure risk reduction.
7. Stratification by sex.

For all Cox models, proportional hazard assumptions were visually checked and confirmed. All analyses were conducted using R (version 4.1.3; R Foundation for Statistical Computing, Austria).

### Patient and public involvement

Patients were not directly involved in the design of this study, but preliminary results were presented to the Exeter Diabetes Patient and Public Involvement Group who advised on the interpretation of these results.

## RESULTS

Amongst n=57,368 SGLT2i and n=111,673 DPP4i/SU comparator arm drug initiations (ESM Figure 1) in those with type 2 diabetes (T2D) and no atherosclerotic cardiovascular disease (ASCVD), heart failure (HF) or chronic kidney disease (CKD), median age was 58.0 years (IQR: 50.4-66.1 years), 57.6% were male, 74.6% were of White ethnicity, and median follow-up was 2.0 years (IQR: 0.9-3.6 years). Overlap weighting using propensity scores resulted in well-balanced covariates (Table 1, ESM Figure 2).

**Figure 2:**
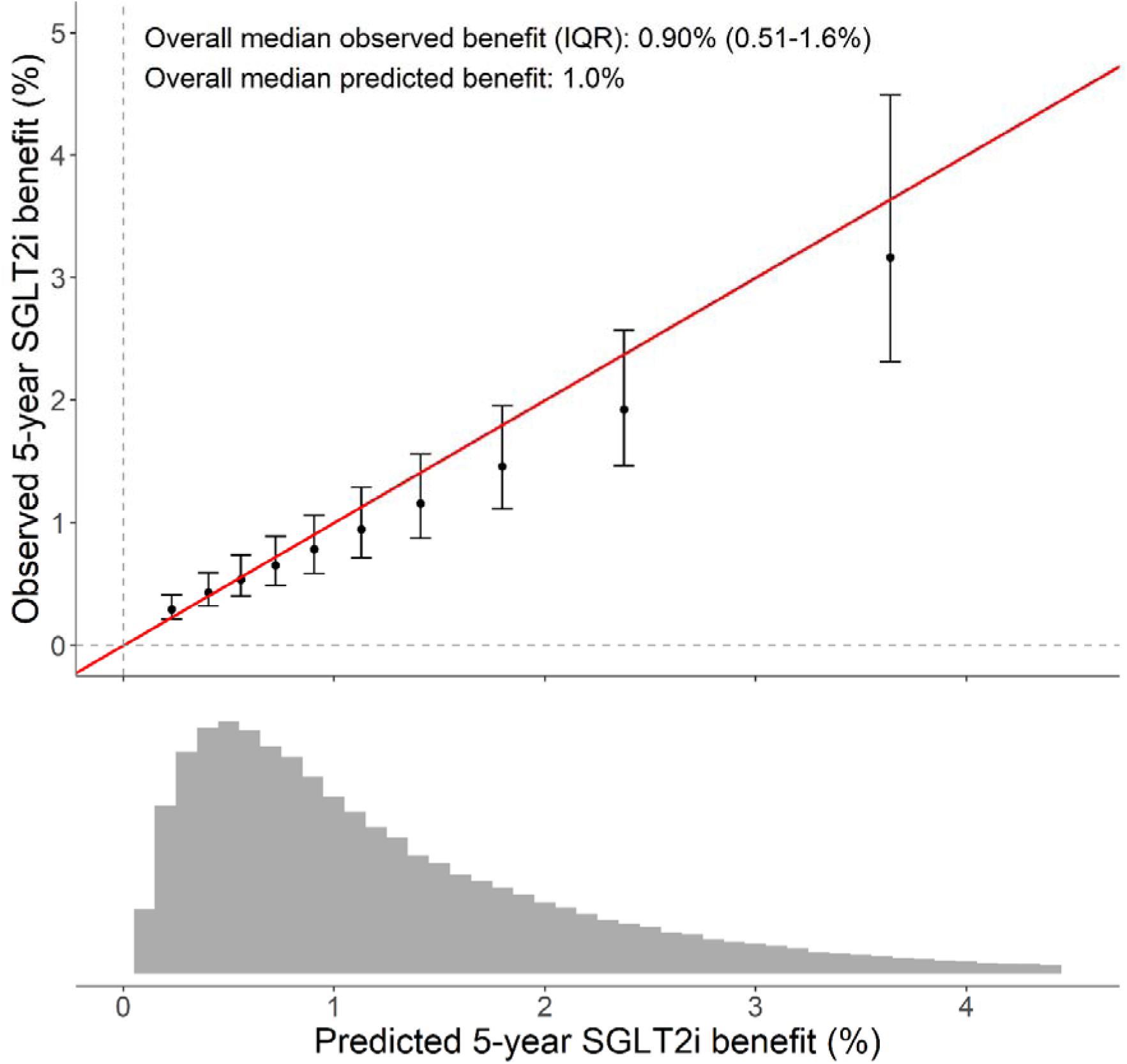
Calibration between 5-year predicted (from SABRE) and observed absolute SGLT2i benefit estimates in the study cohort. Values are medians by decile of predicted benefit, IQRs also shown for observed estimates (n=169,041). Lower plot shows distribution of 5-year predicted benefits (n=164,894; x-axis truncated at 4.5% excluding n=4,147 with predicted benefit >4.5%). SABRE= SGLT2i absolute benefit response model; SGLT2i=SGLT2-inhibitor.

**Table 1:** Baseline characteristics of study cohort at drug (SGLT2-inhibitor/comparator) initiation after overlap weighting. Unweighted characteristics shown in ESM Table 1.

|  | **Comparator**<br>**(DPP4i/SU)** | **SGLT2i** |
| --- | --- | --- |
| **Sex (% male)** | 57.2% | 57.2% |
| **Age at drug initiation (years)** | 57.1 (49.6-65.2) | 57.3 (50.2-64.6) |
| **Diabetes duration (years)** | 6.8 (3.6-11.1) | 6.9 (3.8-11.1) |
| **Ethnicity** |  |  |
| White | 75.4% | 75.4% |
| Asian | 14.1% | 14.1% |
| Black | 4.5% | 4.5% |
| Other | 3.6% | 3.6% |
| Missing | 2.5% | 2.5% |
| **Index of Multiple Deprivation quintile** |  |  |
| 1 (least deprived) | 16.9% | 16.9% |
| 2 | 17.7% | 17.7% |
| 3 | 19.2% | 19.2% |
| 4 | 22.5% | 22.5% |
| 5 (most deprived) | 23.7% | 23.7% |
| **Smoking status** |  |  |
| Non-smoker | 52.8% | 52.8% |
| Active smoker | 16.2% | 16.2% |
| Ex-smoker | 31.1% | 31.1% |
| **Hypertension** | 53.9% | 53.9% |
| **Atrial fibrillation** | 2.3% | 2.3% |
| **Number of hospital admissions in previous year** |  |  |
| 0 | 80.9% | 80.9% |
| 1 | 16.5% | 16.5% |
| 2+ | 2.6% | 2.6% |
| **BMI (kg/m<sup>2</sup>)** | 32.1 (28.2-37.0) | 32.4 (28.9-36.9) |
| **HbA1c (mmol/mol)** | 72.0 (63.0-86.0) | 73.0 (64.0-86.0) |
| **SBP (mmHg)** | 132.0 (124.0-140.0) | 132.0 (124.0-140.0) |
| **Total cholesterol:HDL** | 3.8 (3.1-4.7) | 3.8 (3.1-4.7) |
| Missing | 3.6% | 3.5% |
| **Drug line** |  |  |
| 2 | 36.8% | 36.8% |
| 3 | 38.8% | 38.8% |
| 4 | 17.8% | 17.8% |
| 5+ | 6.7% | 6.7% |
| **Number of other current non-insulin glucose-lowering medications** |  |  |
| 0 | 9.0% | 9.0% |
| 1 | 57.2% | 57.2% |
| 2+ | 33.8% | 33.8% |
| **Current insulin use** | 5.3% | 5.3% |
| **Year of drug initiation** |  |  |
| 2013 | 2.6% | 2.6% |
| 2014 | 8.0% | 8.0% |
| 2015 | 12.9% | 12.9% |
| 2016 | 14.3% | 14.3% |
| 2017 | 16.1% | 16.1% |
| 2018 | 17.6% | 17.6% |
| 2019 | 18.1% | 18.1% |
| 2020 | 10.3% | 10.3% |
| **QDiabetes-Heart Failure 5-year score (%)** | 2.7 (1.5-4.8) | 2.7 (1.5-4.7) |

All values are % or median (interquartile range).
DPP4i=DPP4-inhibitor, SGLT2i=SGLT2i-inhibitor, SU=sulfonylurea.

### SGLT2i initiation is associated with a lower relative risk of new-onset heart failure, irrespective of baseline absolute risk

1.2% (706/57,368) of the SGLT2i initiators and 1.8% (2,052/111,673) of the DPP4i/SU initiators experienced new-onset heart failure during the study period (incidence rates: SGLT2i: 5.60 per 1000 patient-years, DPP4i/SU: 7.78 per 1000 patient-years). Overall, we found a 30% lower relative risk of new-onset heart failure with SGLT2i versus the comparator arm (adjusted and weighted HR: 0.70 [95% CI: 0.63 to 0.78]), concordant with the previous trial meta-analysis for hospitalisation for heart failure in a similar population (HR: 0.63 [95% CI: 0.50 to 0.80]). This lower relative risk was consistent across all sensitivity analyses, including stratification by sex (HR range 0.65-0.72, ESM Figure 3). A consistently lower relative risk with SGLT2i was also seen at all levels of predicted absolute heart failure risk (Figure 1), where we found no evidence that the relative heart failure benefit associated with SGLT2i was modified by baseline HF risk (p=0.82 for treatment arm:baseline HF risk interaction). This was consistent in males and females (interaction p=0.80 [males] and 0.17 [females]).

**Figure 3:**
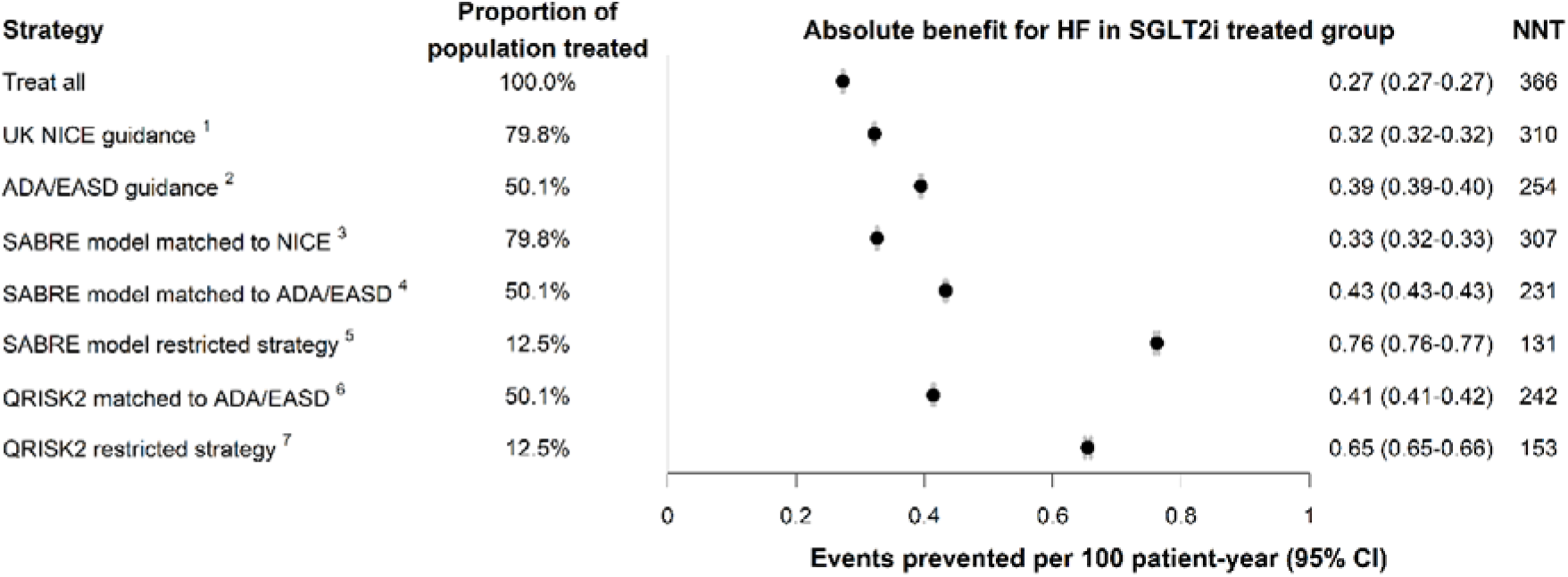
Modelled absolute benefit (primary heart failure (HF) events prevented) in SGLT2i-treated individuals selected by different SGLT2-inhibitor targeting strategies. Based on study population of individuals with type 2 diabetes without atherosclerotic cardiovascular disease, heart failure or chronic kidney disease. Absolute benefit is number of primary HF events prevented per 100 patient-years of treatment in those treated, derived from SABRE model predictions. NNT is number of patients needed-to-treat per year to prevent one HF event. ^1^Treat individuals with QRISK2 10-year score >10%. ^2^Treat individuals aged ≥55 years with 2 or more cardiovascular risk factors, including obesity, hypertension, or dyslipidaemia. ^3^Treat individuals with SABRE predicted 5-year heart failure benefit >0.48%, resulting in recommending SGLT2i to the same proportion of individuals as UK NICE guidelines. ^4^Treat individuals with SABRE predicted heart failure benefit >1.0%, resulting in recommending SGLT2i to the same proportion of individuals as ADA/EASD guidelines. ^5^Treat individuals with SABRE predicted heart failure benefit >2.6%, corresponding to the top 25% centile of predicted heart failure benefit in those recommended SGLT2i by ADA/EASD guidelines. ^6^Treat individuals with 10-year QRISK2 score >19.1%, resulting in recommending SGLT2i to the same proportion of individuals as ADA/EASD guidelines. ^7^Treat individuals with 10-year QRISK2 score >36.3%, corresponding to the top 25% centile of QRISK2 in those recommended SGLT2i by ADA/EASD guidelines. HF=heart failure, SABRE= SGLT2i absolute benefit response model, SGLT2i=SGLT2i-inhibitor.

### The QDiabetes-HF prediction model accurately predicts absolute risk of new-onset heart failure

5-year predictions of absolute heart failure risk were well-calibrated, although risk was slightly under-estimated in the highest risk decile (ESM Figure 4). The C-statistic was 0.72 (95% CI: 0.70 to 0.73), compared to 0.77-0.78 in the original validation^6^, and the Brier score was 0.036 (95% CI: 0.034 to 0.038), suggesting overall good calibration and accuracy.

**Figure 4:**
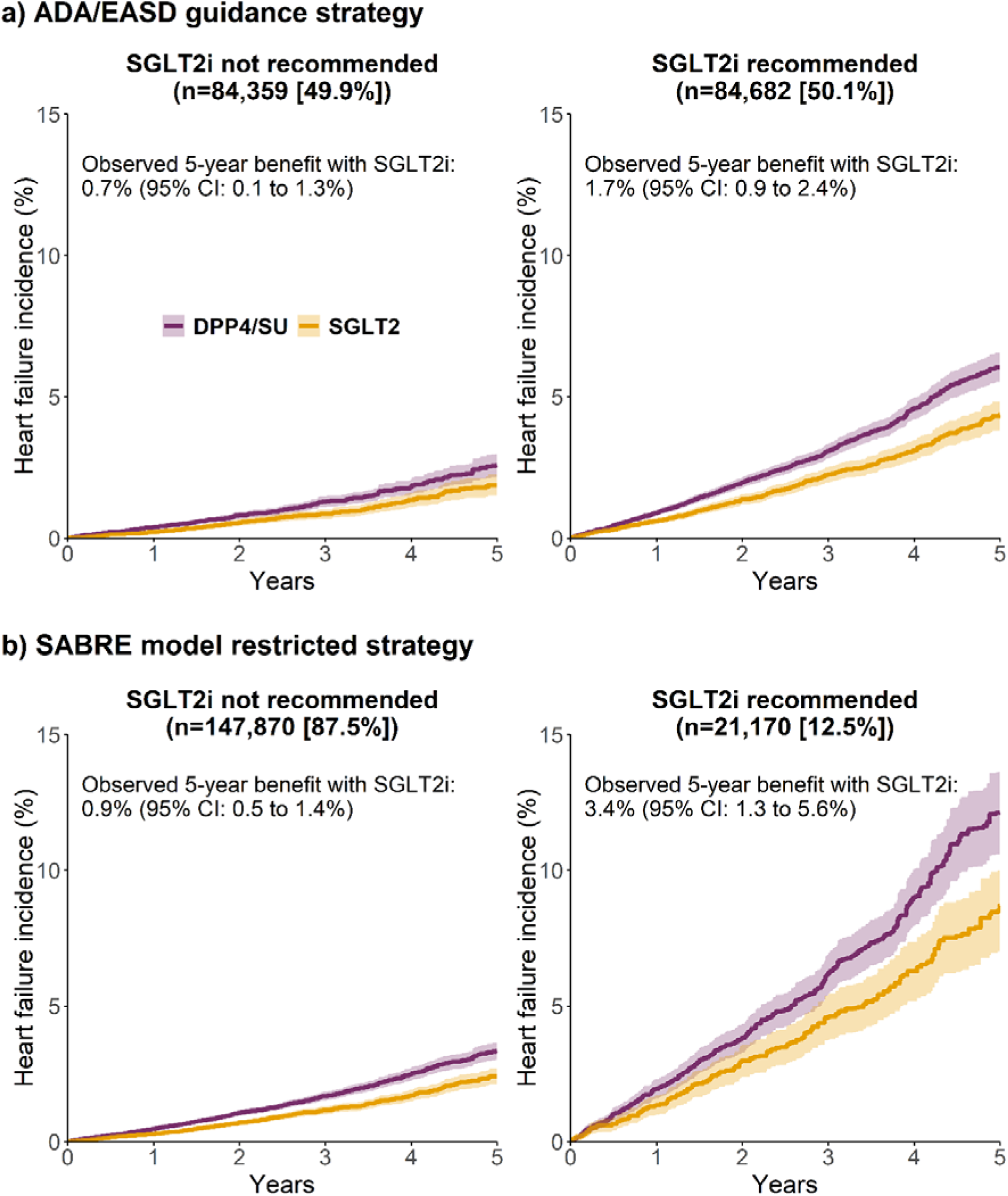
Kaplan-Meier plots of observed heart failure incidence over 5 years for SGLT2-inhibitor treated individuals vs comparator (treated with DPP4-inhibitor/sulfonylurea), with patients stratified based on different SGLT2i treatment strategies: a) ADA/EASD guidance, b) SABRE model ‘restricted’ strategy (predicted absolute heart failure benefit >2.6%). Overlap weighting was used within each stratum to balance SGLT2-inhibitor and comparator groups. Observed benefits are estimated differences in survival at 5 years between the weighted SGLT2-inhibitor and comparator groups. DPP4i=DPP4-inhibitor, SABRE= SGLT2i absolute benefit response model, SGLT2i=SGLT2i-inhibitor, SU=sulfonylurea.

### Absolute heart failure benefit predictions from the SABRE model are accurate in validation

Predicted individual-level 5-year SGLT2i benefits from the SGLT2i absolute benefit response model (SABRE) ranged from <0.1% to 14.1% (median [IQR] 1.0% [0.6-1.8%]; ESM Figure 5). In validation, predicted and observed SGLT2i benefits were well-calibrated, overall and by decile of predicted SGLT2i benefit (Figure 2).

### SABRE allows more precise targeting of SGLT2i treatment for heart failure benefit compared with current guidelines

The expected outcomes of different treatment strategies, in terms of the proportion of those with T2D without ASCVD/HF/CKD who would be SGLT2i-treated, absolute benefit for those treated, and numbers needed-to-treat, are shown in Figure 3. The results show that deploying the SABRE model at a threshold matched to current ADA/EASD guidance (treat 50% of patients; predicted 5-year heart failure benefit >0.48%) results in a lower 1-year NNT (231 vs 254 for ADA/EASD) by more precisely targeting those with greatest predicted heart failure benefit (0.43 [95% CI: 0.43 to 0.43] vs 0.39 [95% CI: 0.39 to 0.40] HF events prevented per 100 patient-years in those treated). In addition, the SABRE model can facilitate a restricted treatment strategy by targeting the top quartile of predicted benefit in those indicated by ADA/EASD guidance (predicted 5-year heart failure benefit >1.0%); reducing 1-year NNT to 131, with 0.65 (95% CI: 0.65 to 0.66) HF events prevented per 100 patient-years in the 12.5% of those (with T2D without ASCVD/HF/CKD) treated.

SABRE is predicted to perform marginally better than QRISK2 when used to target SGLT2i to the same proportion of those with T2D without ASCVD/HF/CKD: treating 50% using QRISK2 >19.1% is predicted to prevent 0.41 (95% CI: 0.41 to 0.42) HF events per 100 patient-years in those treated compared to 0.43 (95% CI: 0.43 to 0.43) HF events per 100 patient-years for SABRE (1-year NNT for QRISK2: 242 vs SABRE: 231); and treating 12.5% of the population using QRISK2 >36.3% is predicted to prevent 0.65 (95% CI: 0.65 to 0.66) HF events per 100 patient-years in those treated compared to 0.76 (95% CI: 0.76 to 0.77) HF events per 100 patient-years for SABRE (1-year NNT for QRISK2: 131 vs SABRE: 153). The current UK guidance strategy (QRISK2 >10%) recommends treatment for most individuals (79.8% of those with T2D without ASCVD/HF/CKD) with a higher 1-year NNT of 366.

Figure 4 shows observed 5-year incidence of heart failure with treatment initiation defined by ADA/EASD guidance and the SABRE restricted strategy, with outcomes for the other treatment strategies shown in ESM Figure 6. The absolute observed heart failure benefit with SGLT2i versus the comparator arm was greater in those receiving treatment with the SABRE restricted strategy (3.4% benefit, 95% CI: 1.3 to 5.6%) than the ADA/EASD guidance strategy (1.7% benefit, 95% CI: 0.9 to 2.4%). Despite the higher proportion of individuals not indicated for treatment under the SABRE strategy, observed outcomes in those not recommended for treatment were similar (SABRE: 0.9%, ADA/EASD: 0.7%). The QRISK2 ‘restricted’ threshold of >36.3% led to a similarly greater observed heart failure benefit in those treated (2.6% benefit [95% CI: 0.5 to 4.7%], treat 12.5%; ESM Figure 6e).

## Discussion

### Principle findings

We develop and validate a SGLT2i heart failure benefit prediction model (SABRE) allowing accurate identification of individuals who would benefit most from SGLT2i for primary heart failure prevention. The model is applicable to people with type 2 diabetes without atherosclerotic cardiovascular disease (ASCVD), heart failure (HF) or chronic kidney disease (CKD), which represent the majority of the T2D population in the UK^4^ ^27^. Importantly, we establish that although there is no evidence of variation in the relative SGLT2i benefit for heart failure in our large population-representative T2D population without ASCVD/HF/CKD, there is substantial variation in absolute benefit, and this allows for prioritisation of SGLT2i to those with greatest predicted absolute benefit. In our UK cohort, SABRE enabled more precise targeting than existing SGLT2i targeting strategies from ADA/EASD ^2^ and NICE ^3^.

Direct low-cost implementation of SABRE is possible as the model combines RCT results with an established risk prediction model based on routine clinical features which can easily be updated for deployment in other settings.

### Strengths and weaknesses compared to existing studies

Our observational finding of an overall 30% lower risk of new-onset heart failure in those with T2D and no pre-existing ASCVD with SGLT2i (HR: 0.70 [95% CI: 0.63 to 0.78]) is consistent with previous RCT meta-analysis ^1^ ^28^ (HR: 0.63 [95% CI: 0.49-0.50 to 0.80] for first hospitalisation for heart failure in both meta-analyses). Importantly, we show this average estimate appears constant across the range of continuous baseline heart failure risk seen in our large multi-ethnic and population-representative T2D population. This consistent relative benefit has been previously suggested across subgroups defined by baseline heart failure risk in RCT data ^29^ and supports potential generalisability of the SABRE model to other populations in which heart failure risk patterns may be different to the UK. A recent study has suggested that whilst ASCVD is decreasing in those with T2D, heart failure incidence has now plateaued or even started to increase, especially in younger patients ^30^, supporting heart failure as a relevant treatment target.

SABRE predicted that treating our entire study population with SGLT2i would prevent 0.27 (95% CI: 0.27 to 0.27) HF events per 100 patient-years; less than the 0.39 (95% CI: 0.32 to 0.47) events per 100 patient-years reported in an existing meta-analysis of observational studies in those with T2D and no ASCVD ^31^, likely reflecting lower baseline heart failure risk in our study population. The same meta-analysis showed a much greater benefit in those with T2D and ASCVD (for whom SGLT2i are currently recommended) of 1.17 (95% CI: 0.78 to 1.55) HF events prevented per 100 patient-years ^31^. In our study, targeting SGLT2i to the 12.5% of those with T2D without ASCVD/HF/CKD with greatest predicted benefit increased the benefit to 0.76 (95% CI: 0.76 to 0.77) HF events prevented per 100 patient-years, still below the benefit seen in those with T2D and ASCVD.

### Strengths and weaknesses

Strengths of this study include the use of existing evidence on SGLT2i benefit from clinical trial meta-analysis and a validated risk prediction model to develop the SABRE model. The final model was then validated in a large, population-representative dataset. We focused on individuals with T2D who do not have ASCVD, HF, or CKD, as evidence-based guidance already recommends initiation of SGLT2i for individuals with these conditions. In contrast to the original RCTs included in the trial meta-analysis, in which up to 25% of patients had CKD and/or macroalbuminuria, our study population excluded those with chronic kidney disease (stage 3a-5) or macroalbuminuria as they are already indicated for SGLT2i under current guidance. Our finding of a similar relative risk reduction with SGLT2i for HF as the meta-analysis therefore suggests that the presence of CKD and/or macroalbuminuria does not substantially affect the relative benefit of SGLT2i for heart failure.

A limitation of our study is that both the study population used here to validate SABRE, and those used in the development of QDiabetes-HF, were of majority White ethnicity and aged <84 years. The need for ethnicity-specific models should be evaluated carefully in future work. There is limited evidence from RCTs on the relative effectiveness of SGLT2i for cardiovascular outcomes across different ethnicities ^32^ and particular for primary heart failure prevention in T2D, and we did not have large enough numbers to assess this in our study. A further limitation is the accuracy of the QDiabetes-HF model used in SABRE to predict heart failure risk: the C-statistic of 0.72 in our study population is slightly lower than that achieved in the original validation 0.77-0.78^6^, which may reflect temporal changes since model development and/or differences between the development cohort and our study cohort.

Routinely available risk factors for heart failure including chronic obstructive pulmonary disease, obstructive sleep apnoea, and excessive alcohol use ^33^ are not included in QDiabetes-Heart Failure and could improve accuracy of predictions. An enhanced heart failure prediction model might provide a greater performance benefit over QRISK2 (a lower number-needed-to-treat) for targeting SGLT2i for heart failure outcomes than the similar performance observed in this study. SABRE could easily be updated with new heart failure risk prediction models if/when these are available.

Another limitation of our study is that validation was performed using observational data of patients receiving routine care (i.e. not randomised to treatment). This introduces the possibility of treatment bias which will not have been fully accounted for in our analysis.

However, the concordance between previous RCT results and our estimates of SGLT2i-associated heart failure benefit suggests that any remaining bias is unlikely to explain our results. We have not specifically examined differences in SGLT2i dose or adherence, which may impact heart failure outcomes. However, our data reflects real-world clinical practice and patient behaviours and so represents typical UK prescribing patterns and medication use.

### Potential implications for clinicians or policymakers

The SABRE model provides new individual-level risk estimation to support targeting of SGLT2i within the majority of the T2D population, who do not have ASCVD, HF or CKD, to those who would benefit the most for heart failure prevention. This group of patients represent a significant and increasing population for whom current preventative guidance is largely based on ASCVD outcomes rather than heart failure, and is inconsistent across the US, UK and Europe ^2^ ^3^.

Although our study focused on heart failure, an important point is that those at highest risk of heart failure are generally also at highest risk of other CVD outcomes ^34^, and so prioritising treatment based on heart failure risk will largely align with a broader CVD-targeted strategy. In the UK, our analysis supports the use of QRISK2, a CVD risk model which is established in UK clinical practice, as a pragmatic alternative to the QDiabetes-HF heart failure-specific model used in this study. Currently, QDiabetes-HF is available as an online risk calculator ^35^ but is not routinely used in UK primary care. We show use of QRISK2 is likely to lead to only a modest loss in precision in terms of those targeted (for example, 1-year number needed to treat for heart failure: 153 for QRISK2 vs 131 for SABRE model when 12.5% of the population is treated; Figure 3). This finding could support a cost-saving refinement of recently updated UK guidance which recommends considering SGLT2i for >90% of those with T2D and no cardiovascular disease ^4^. A higher 10-year QRISK2 >19.1% would target SGLT2i to 50% of those with T2D and no ASCVD/HF/CKD, a similar proportion to ADA/EASD guidance.

Although we developed and validated SABRE in UK data, simple deployment is possible in other populations. Before deployment, accuracy assessment using setting-specific data, or replacement of the underlying absolute risk model with one that has previously been locally validated, will be required. An advantage of the model is that the number of patients indicated for treatment can be flexibly adjusted by changing the benefit threshold used, to align with local or national cost considerations. Of note, even in those with substantial predicted heart failure benefit with SGLT2i, individual treatment decisions should carefully consider the potential side effects of SGLT2i, especially in frail or elderly individuals ^36^, as well as other benefits including glycaemic response ^37^ and weight improvement.

### Unanswered questions and future research

In addition to heart failure benefits, SGLT2i have demonstrated significant benefits for kidney protection ^1^ which we have not taken into consideration. Although heart failure and chronic kidney disease share many risk factors ^38^, there are likely to be patients who would benefit substantially from SGLT2i treatment for kidney protection but are not identified by the SABRE model as having high predicted heart failure benefit. Future work will seek to identify and describe these patients.

Our analysis did not incorporate GLP-1 receptor agonists (GLP-1RA), for which next generation agents have recently been shown to reduce the risk of atherosclerotic cardiovascular disease (ASCVD) and heart failure (the latter to a lesser degree than SGLT2i)^39^. In ADA/EASD guidance, GLP-1RA are recommended as an alternative to SGLT2i for those with T2D at high CVD risk ^2^, whereas in the UK they are currently only recommended for limited subset of those with T2D (predominantly those with obesity) ^3^. As both drug classes have also been shown to reduce heart failure and cardiovascular disease in individuals without diabetes ^39^ ^40^, future studies applying our research framework to understand the absolute benefits in both diabetes and non-diabetes populations would be of considerable interest.

## Conclusion

We have developed and validated the SABRE model to predict the absolute benefit for heart failure prevention of initiating SGLT2i for people with type 2 diabetes without ASCVD, heart failure or chronic kidney disease. The model framework, based on RCT evidence and validated clinical prediction models, means it can be quickly adapted for deployment in multiple countries. Deployment could support a more targeted approach to prescribing of SGLT2i in people with type 2 diabetes to maximise population-level benefit.

## Supporting information

ESM

TRIPOD-AI checklist

## Data Availability

Access to CPRD data is subject to protocol approval via CPRD's research data governance process (https://cprd.com/data-access).

## Contributors

The study concept and design were conceived and developed by KGY, BMS and JMD. KGY, RH and JMD undertook the analysis, with support from BMS. APM reviewed the clinical codes. All authors provided support for the analysis and interpretation of results, critically revised the manuscript, and approved the final manuscript. KGY and JMD attests that all listed authors meet authorship criteria and that no others meeting the criteria have been omitted. All authors had final responsibility for the decision to submit for publication.

## Transparency statement

The lead author (KGY) is the guarantor of this manuscript and affirms that the manuscript is an honest, accurate, and transparent account of the study being reported; that no important aspects of the study have been omitted; and that any discrepancies from the study as originally planned have been explained. The study protocol was no pre-registered.

## Ethics approval

This study was approved by the Clinical Practice Research Datalink independent scientific advisory committee (protocol 22_002000). CPRD also has ethical approval from the Health Research Authority to support research using anonymised patient data (research ethics committee reference 21/EM/0265). Individual patient consent was not required as all data were deidentified.

## Data sharing

Access to CPRD data is subject to protocol approval via CPRD’s research data governance process (https://cprd.com/data-access). Code for initial cohort preparation is available at: https://github.com/Exeter-Diabetes/CPRD-Cohort-scripts/tree/Oct2020-download/03-Treatment-response-(MASTERMIND) and analysis code is available at https://github.com/Exeter-Diabetes/CPRD-Katie-SGLT2-HF. The QDiabetes-Heart Failure algorithm is available as open-source software under the GNU Aferro General Public Licence, version 3 from https://qdiabetes.org/heart-failure/index.php.

