## Supplementary material for "Precision prescribing of SGLT2-inhibitors in people with type 2 diabetes for primary prevention of heart failure: model development and validation study": ESM

### ESM Methods

#### Variable definitions

All codelists for defining baseline variables used for QDiabetes-Heart Failure, QRISK2, regression model adjustment/propensity scores, as well as heart failure and cardiovascular outcomes, can be found here: https://github.com/Exeter-Diabetes/CPRD-Codelists/tree/pre-2024. In addition, this repository has algorithms for defining date of birth, diabetes type, diabetes duration, ethnicity, smoking status, chronic kidney disease status, as well as cleaning rules for diagnoses, prescriptions and biomarkers.

Additional information on variables not fully defined by the above:

- Cardiovascular disease was present at baseline if there was any record of angina, ischaemic heart disease, myocardial infarction, peripheral arterial disease, revascularisation, stroke or transient ischaemic attack in primary or secondary care data prior to baseline.
- Heart failure was present at baseline if there was any record of heart failure in primary or secondary care data prior to baseline.
- Chronic kidney disease was present at baseline if there was any record of chronic kidney disease stage 3a-5 or end stage renal disease or macroalbuminuria (urinary albumin:creatinine >30mg/dl) in primary or secondary care data prior to baseline.
- Ethnicity was coded as 9-category QRISK2 ethnicity (NB: the model treats missing ethnicity as White).
- Smoking status was coded as 5-category QRISK2 smoking status.
- For defining current medication use, gaps of <6 months between subsequent prescriptions for the same drug class were ignored (represented continuous treatment) as per our previously published protocol (1).
- Drug initiations within 3 months of patient registration at the primary care practice were excluded as they may not represent new initiations.
- Number of glucose-lowering drug classes ever prescribed was categorised as 1, 2, 3, 4 or 5+ distinct glucose-lowering drug classes (including the current drug being initiated).
- Number of other current non-insulin glucose-lowering medications (metformin, sulfonylurea, DPP4-inhibitor, SGLT2-inhibitors, thiazolidinedione or GLP-1 receptor agonist; not including drug [SGLT2i/DPP4i/SU] being initiated) was categorised as 0, 1 or 2+.
- Number of emergency inpatient hospital admissions in the previous year (admissions recorded in Hospital Episode Statistics Admitted Patient Care [HES APC] data with an admission method other than ‘elective’) was categorised as 0, 1-2 or 3+.
- Deprivation scores: 2015 English Index of Multiple Deprivation (IMD) decile scores were used for propensity scores and hazard ratio adjustment. Townsend Deprivation Scores (TDS) were required for QDiabetes-Heart Failure and QRISK2 risk scores. To estimate TDS scores from IMD, we used 2015 IMD deciles and 2011 TDS scores for all Lower Super Output Areas (LSOAs) in England, and found the median 2011 TDS corresponding to each IMD decile score.
- ‘Current blood pressure medication’ (used in QRISK2) was defined as at least 2 prescriptions for thiazide, beta-blocker, calcium channel blocker, or angiotensin converting enzyme inhibitor, with at least one in the last 28 days prior to baseline date as per (2).
- Obesity (used for ADA-EASD high cardiorenal risk definition) was defined as BMI >30 $kg/m^2$.

- Hypertension (used for adjustment/weighting and ADA-EASD high cardiorenal risk definition) was defined as a previous diagnosis of hypertension in primary care data or SBP>140 mmHg or DBP>90 mmHg (SBP and DBP assumed to be normal if missing).
- Dyslipidaemia (used for ADA-EASD high cardiorenal risk definition) was defined as current statin treatment (as above, gaps of <6 months between subsequent prescriptions were ignored) or total cholesterol >5 mmol/L.
- Albuminuria (used for ADA-EASD high cardiorenal risk definition) was defined as urinary albumin:creatinine ratio of >3 mg/mmol (assumed to be normal if missing).
- Total cholesterol:HDL ratio was calculated from separate total cholesterol and HDL codes and imputed from age, sex, ethnicity and smoking status where missing as per the QRISK2 and QDiabetes-Heart Failure score algorithms.
- Patients with biomarker values outside the range allowed by QRISK2 and QDiabetes-Heart Failure were excluded (HbA1c: <40 or >150 mmol/mol, cholesterol:HDL <1 or >11 mmol/L, SBP <70 or >210 mmHg).
- All biomarkers (HbA1c, total cholesterol, cholesterol:HDL, SBP, DBP, BMI, urinary albumin:creatinine ratio) were within the 2 years prior or up to 7 days after baseline date. HbA1c measurements were taken a median of 15 days before drug initiation, BMI 27 days, total cholesterol 32 days, HDL 35 days, SBP 13 days, DBP 13 days, urinary albumin:creatinine ratio 123 days; 83.6% of biomarker measurements were within 6 months of drug initiation.

**QRISK2 variables**

QRISK2, like QDiabetes-Heart Failure, includes age, sex, ethnicity (9 groups derived from the 16 UK census categories: White, Indian, Pakistani, Bangladeshi, Black Caribbean, Black African, Chinese, Other), deprivation, smoking status, diabetes type, atrial fibrillation, chronic kidney disease stage 4 or 5, total cholesterol:HDL ratio, systolic blood pressure (SBP) and BMI as predictors. QRISK2 additionally includes blood pressure medication use, premature cardiovascular disease in a first degree relative, and rheumatoid arthritis. Unlike QDiabetes-Heart Failure, QRISK2 is not specifically designed for people with diabetes. It includes presence of diabetes as a predictor but does not account for diabetes duration or HbA1c. Additionally, QRISK2 does not include a history of heart attack, angina, or stroke since it was developed to assess the risk of primary cardiovascular disease in individuals without a prior diagnosis.

**ESM Tables**

**ESM Table 1: Baseline characteristics of study cohort at drug (SGLT2-inhibitor/comparator) initiation (no weighting).**

| | **Comparator (DPP4i/SU) (N=111,673; N=68,708 [61.5%] DPP4i, N=42,965 [38.5%] SU)** | **SGLT2i (N= 57,368)** |
|---|---|---|
| **Sex (% male)** | 64,473 (57.7%) | 33,041 (57.6%) |
| **Age at drug initiation (years)** | 58.6 (50.6-67.0) | 57.1 (50.2-64.2) |
| **Diabetes duration (years)** | 6.1 (3.1-10.0) | 8.2 (4.6-12.5) |
| **Ethnicity** | | |
| White | 82,817 (74.2%) | 43,346 (75.6%) |
| Asian | 16,129 (14.4%) | 8,194 (14.3%) |
| Black | 5,987 (5.4%) | 2,419 (4.2%) |
| Other | 4,214 (3.8%) | 1,967 (3.4%) |
| Missing | 2,526 (2.3%) | 1,442 (2.5%) |
| **Index of Multiple Deprivation quintile** | | |
| 1 (least deprived) | 18,200 (16.3%) | 9,961 (17.4%) |
| 2 | 19,744 (17.7%) | 10,191 (17.8%) |
| 3 | 21,136 (18.9%) | 11,016 (19.2%) |
| 4 | 25,491 (22.8%) | 12,840 (22.4%) |
| 5 (most deprived) | 27,102 (24.3%) | 13,360 (23.3%) |
| **Smoking status** | | |
| Non-smoker | 58,224 (52.1%) | 30,599 (53.3%) |
| Active smoker | 18,836 (16.9%) | 8,905 (15.5%) |
| Ex-smoker | 34,613 (31.0%) | 17,864 (31.1%) |
| **Hypertension** | 59,321 (53.1%) | 31,526 (55.0%) |
| **Atrial fibrillation** | 2,945 (2.6%) | 1,281 (2.2%) |
| **Number of hospital admissions in previous year** | | |
| 0 | 88,697 (79.4%) | 46,871 (81.7%) |
| 1 | 19,301 (17.3%) | 9,158 (16.0%) |
| 2+ | 3,675 (3.3%) | 1,339 (2.3%) |
| **BMI (kg/m2)** | 31.0 (27.4-35.5) | 32.7 (29.0-37.2) |
| **HbA1c (mmol/mol)** | 71.0 (62.0-85.0) | 74.0 (65.0-87.0) |
| **SBP (mmHg)** | 132.0 (123.0-140.0) | 132.0 (124.0-140.0) |
| **Toatl cholesterol:HDL** | 3.8 (3.1-4.7) | 3.8 (3.1-4.6) |
| Missing | 4604 (4.1%) | 2015 (3.5%) |
| **Drug line** | | |
| 2 | 62,881 (56.3%) | 13,474 (23.5%) |
| 3 | 37,750 (33.8%) | 17,439 (30.4%) |
| 4 | 8,675 (7.8%) | 15,996 (27.9%) |
| 5+ | 2,367 (2.1%) | 10,459 (18.2%) |
| **Number of other current non-insulin glucose-lowering medications** | | |
| 0 | 12,453 (11.2%) | 4,235 (7.4%) |
| 1 | 73,741 (66.0%) | 27,181 (47.4%) |
| 2+ | 25,479 (22.8%) | 25,952 (45.2%) |
| **Current insulin use** | 2,685 (2.4%) | 6,051 (10.5%) |
| **Year of drug initiation** | | |
| 2013 | 16,028 (14.4%) | 862 (1.5%) |
| 2014 | 15,233 (13.6%) | 3,870 (6.7%) |
| 2015 | 16,069 (14.4%) | 7,124 (12.4%) |
| 2016 | 15,943 (14.3%) | 7,822 (13.6%) |

| | | |
|---|---|---|
| 2017 | 15,351 (13.7%) | 8,974 (15.6%) |
| 2018 | 14,423 (12.9%) | 10,411 (18.1%) |
| 2019 | 12,092 (10.8%) | 11,688 (20.4%) |
| 2020 | 6,534 (5.9%) | 6,617 (11.5%) |
| **QDiabetes-Heart Failure 5-year score (%)** | 2.7 (1.5-5.0) | 2.8 (1.6-4.8) |

All values are n (%) or median (interquartile range).

DPP4i=DPP4-inhibitor, SGLT2i=SGLT2i-inhibitor, SU=sulfonylurea.

**ESM Figures**

**ESM Figure 1: Flow diagram of inclusion for the study cohort**

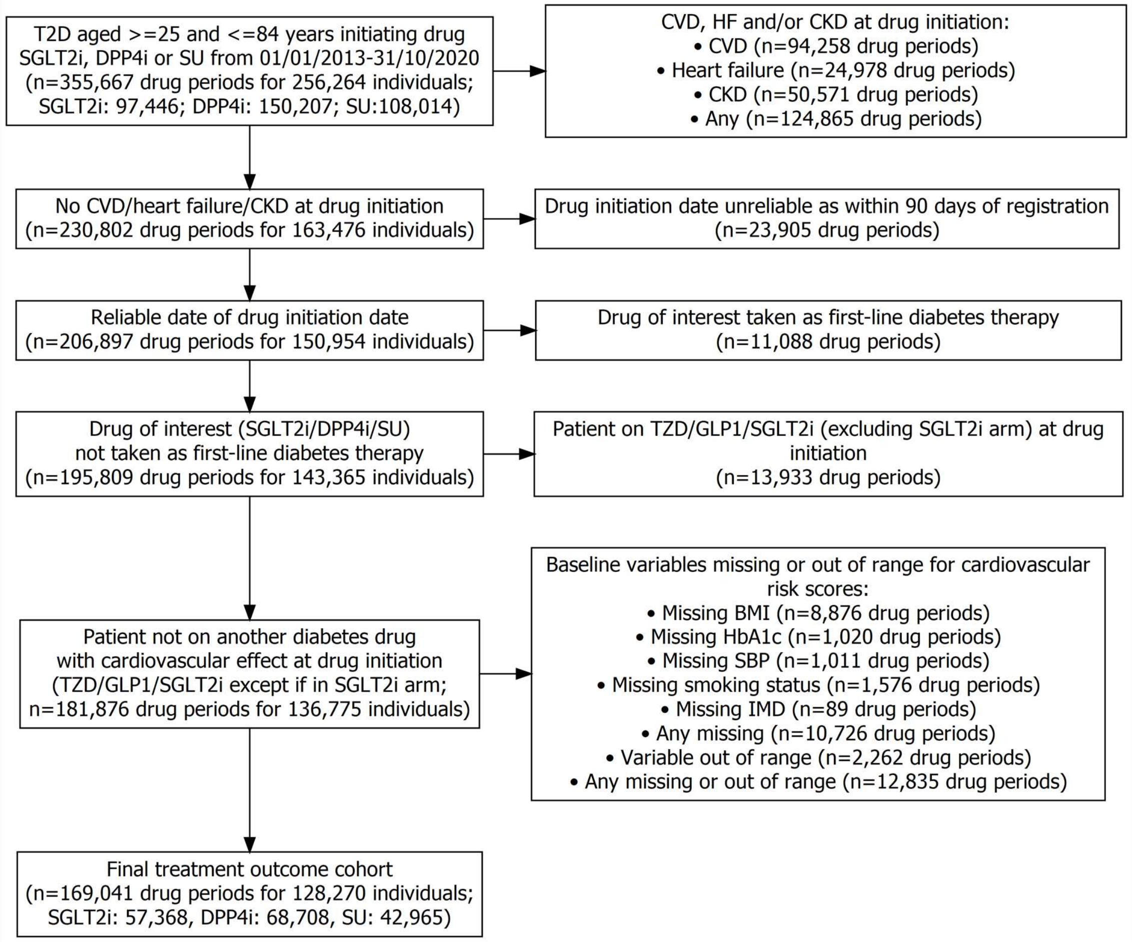


CKD=chronic kidney disease, CVD=cardiovascular disease, DPP4i=DPP4-inhibitor, GLP1=GLP1 receptor agonist, HF=heart failure, SGLT2i=SGLT2i-inhibitor, SU=sulfonylurea, T2D=type 2 diabetes, TZD=thiazolidinedione.

**ESM Figure 2: Love plot of covariate balance in the SGLT2-inhibitor and comparator (DPP4-inhibitor/sulfonylurea; reference group) groups of the treatment outcome cohort before and after overlap weighting.**

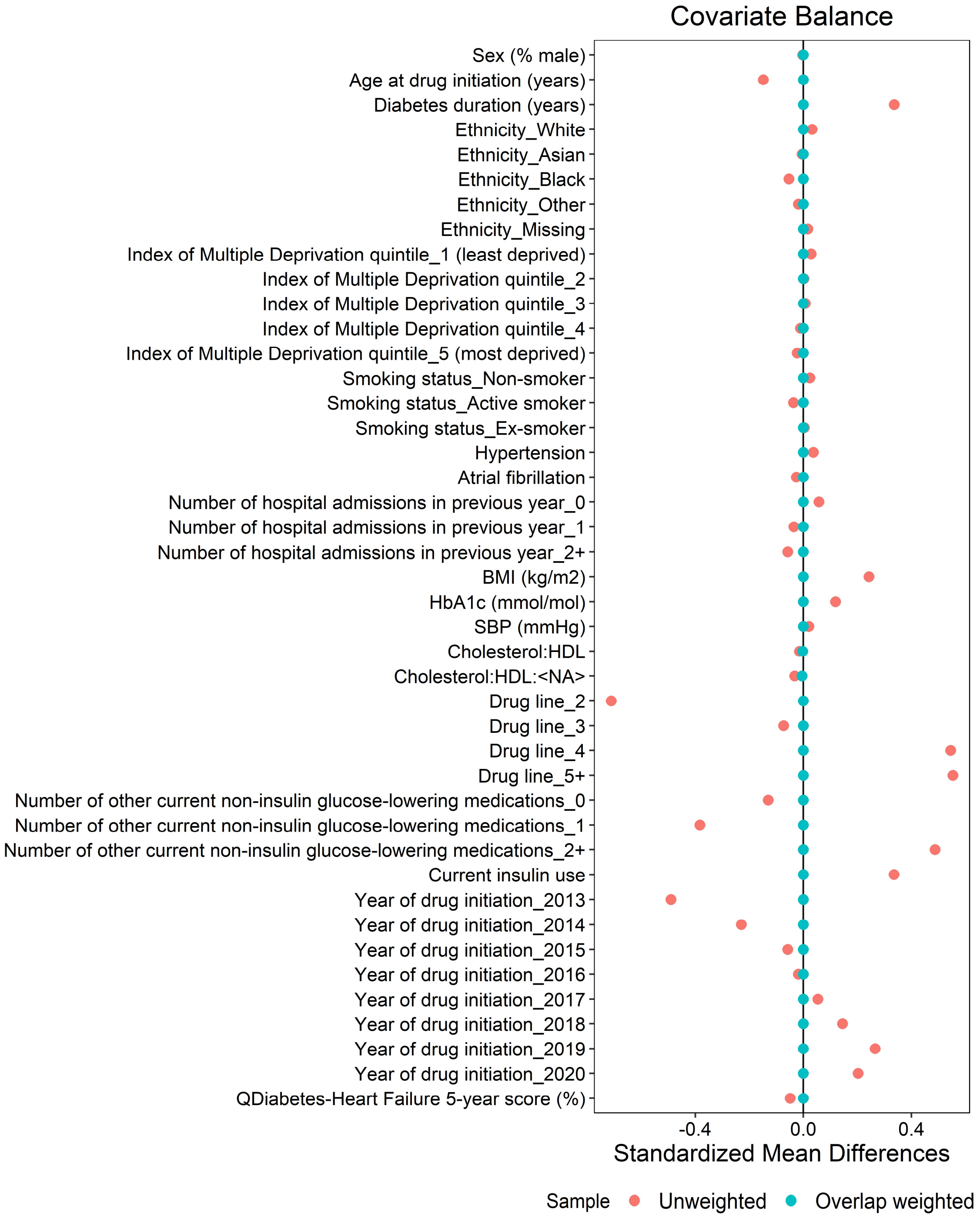

**ESM Figure 3: Hazard ratio estimates for new-onset heart failure for SGLT2-inhibitors vs comparator (DPP4-inhibitors/sulfonylurea) in the study cohort compared to trial meta-analysis.** All models are adjusted with overlap weighting using propensity scores unless otherwise stated (see 'Weighting' section of Methods for variables used for adjustment and propensity scores).

| | SGLT2i Events | IR | Comparator Events | IR | Hazard ratio (95% CI) |
|---|---|---|---|---|---|
| Primary analysis: adjusted with overlap weighting | 706/57368 | 5.60 | 2052/111673 | 7.78 | 0.70 (0.63-0.78) |
| SU-only comparator group | 706/57368 | 5.60 | 826/42965 | 7.69 | 0.69 (0.60-0.80) |
| DPP4i-only comparator group | 706/57368 | 5.60 | 1226/68708 | 7.84 | 0.70 (0.62-0.79) |
| Unadjusted with overlap weighting | 706/57368 | 5.60 | 2052/111673 | 7.78 | 0.68 (0.61-0.76) |
| Adjusted with no weighting | 706/57368 | 5.60 | 2052/111673 | 7.78 | 0.70 (0.63-0.78) |
| Adjusted with IPTW | 706/57368 | 5.60 | 2052/111673 | 7.78 | 0.67 (0.59-0.76) |
| More specific HF definition[1] | 147/57368 | 1.16 | 440/111673 | 1.65 | 0.65 (0.51-0.84) |
| Second-line intention-to-treat[2] | 105/11537 | 3.97 | 1155/52767 | 6.54 | 0.73 (0.59-0.90) |
| Second-line per-protocol[2] | 44/11537 | 2.88 | 470/52767 | 5.32 | 0.61 (0.43-0.84) |
| Death as a competing risk[3] | 706/57368 | 5.60 | 2052/111673 | 7.78 | 0.71 (0.64-0.79) |
| Insulin-treated subgroup only | 151/6051 | 9.57 | 99/2685 | 14.95 | 0.67 (0.49-0.93) |
| Males only | 429/33041 | 5.82 | 1207/64473 | 7.77 | 0.68 (0.59-0.79) |
| Females only | 277/24327 | 5.28 | 845/47200 | 7.79 | 0.72 (0.60-0.86) |
| Trial meta-analysis | | | | | 0.63 (0.50-0.80) |

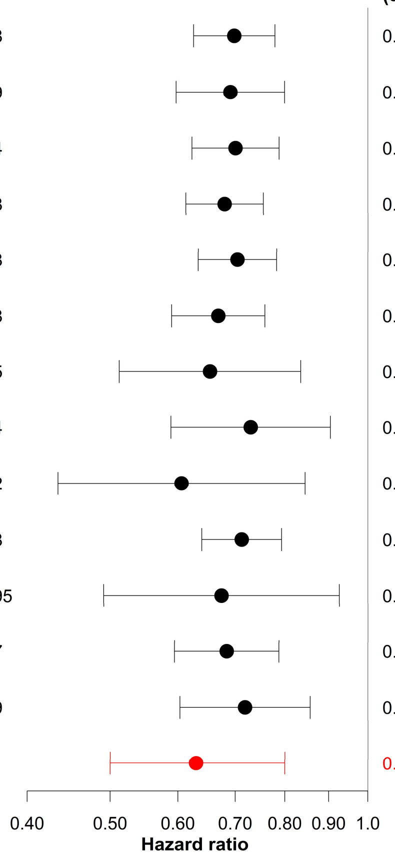


DPP4i: DPP4-inhibitor, HF: heart failure, IPTW: inverse probability of treatment weighting, IR: incidence rate per 1000 patient years, SGLT2i: SGLT2-inhibitor, SU: sulfonylurea.

[1]Outcome includes death or hospitalisation with HF as the primary cause only (main analyses: any code for HF in primary or secondary care or death causes included).

[2]Patients on SGLT2i/DPP4i/SU second-line after metformin. Intention-to-treat: not censored at glucose-lowering treatment change; per-protocol: censored at earliest glucose-lowering treatment change (main analysis: censored if initiate one of the other study drugs, GLP1-receptor agonist or thiazolidinedione).

[3]Death from non-heart failure causes: SGLT2i IR: 5.55, Comparator IR: 9.72. Subdistribution hazard ratio reported as hazard ratio.

**ESM Figure 4: Predicted 5-year absolute risk of new-onset heart failure (from QDiabetes-Heart Failure; median per decile) vs observed estimates for DPP4-inhibitor/sulfonylurea treatment arm (n=111,673).** C-statistic: 0.72 (95% CI: 0.70 to 0.73); Brier score: 0.036 (95% CI: 0.034 to 0.038).

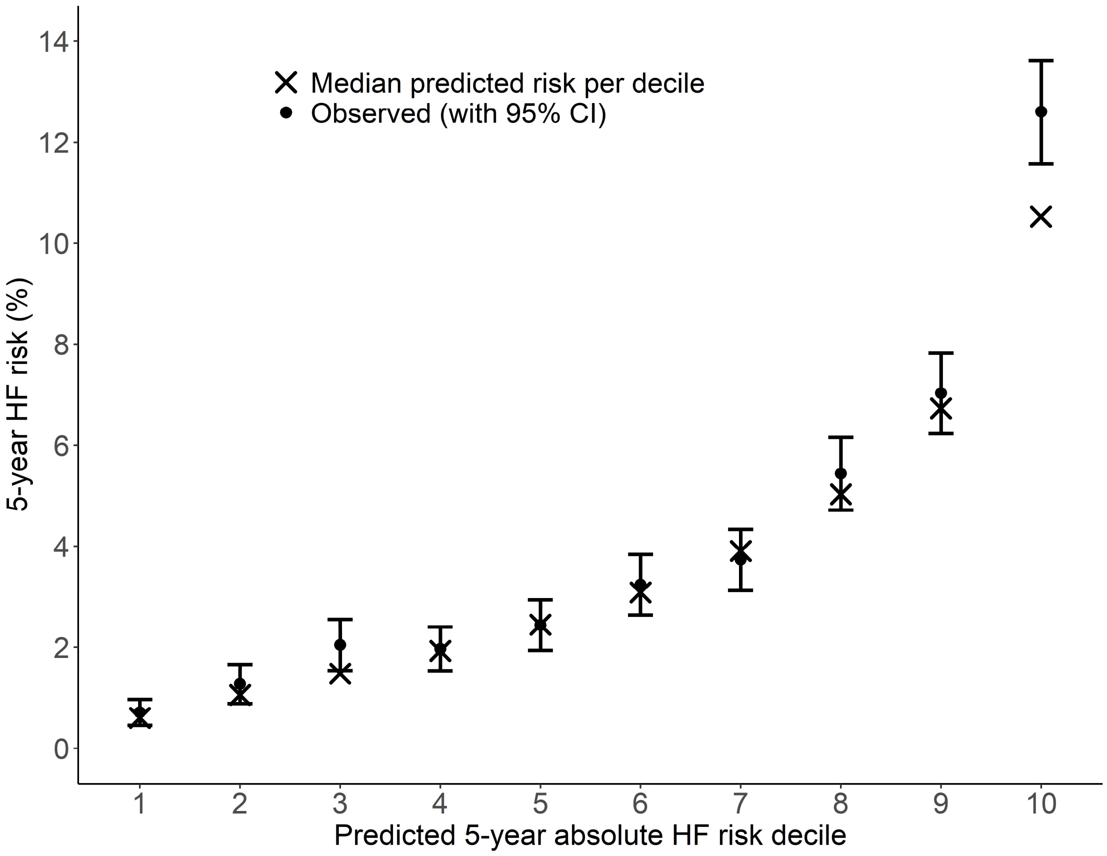


HF=heart failure.

**ESM Figure 5: Distribution of SABRE-predicted absolute SGLT2-inhibitor benefit on 5-year risk of new onset heart failure in the study cohort (n=169,041).**

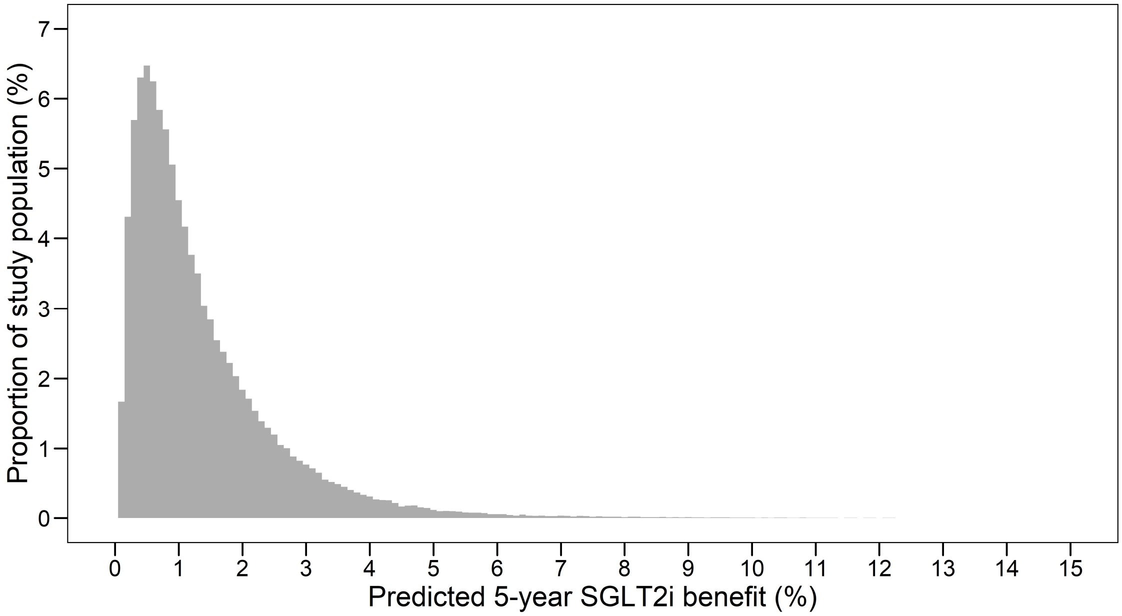


SABRE= SGLT2i absolute benefit response model, SGLT2i=SGLT2i-inhibitor

**ESM Figure 6: Kaplan-Meier plots of observed heart failure incidence over 5 years for SGLT2-inhibitor treated individuals vs comparator (treated with DPP4-inhibitor/sulfonylurea), with patients stratified based on different SGLT2i treatment strategies: a) Current UK NICE guidance (10-year QRISK2 >10%), b) SABRE model matched to NICE (matched to proportions recommended by NICE guidance; predicted absolute heart failure benefit >0.48%), c) SABRE model matched to ADA/EASD (matched to proportions recommended by ADA/EASD guidance; predicted absolute heart failure benefit >1.0%), d) QRISK2 matched to ADA/EASD (matched to proportions recommended by ADA/EASD guidance; 10-year QRISK2 >19.1%), e) QRISK2 'restricted' strategy (10-year QRISK2 >36.3%).** Overlap weighting was used within each stratum to balance SGLT2-inhibitor and comparator groups. Observed benefits are estimated differences in survival at 5 years between the weighted SGLT2-inhibitor and comparator groups.

### a) UK NICE guidance strategy

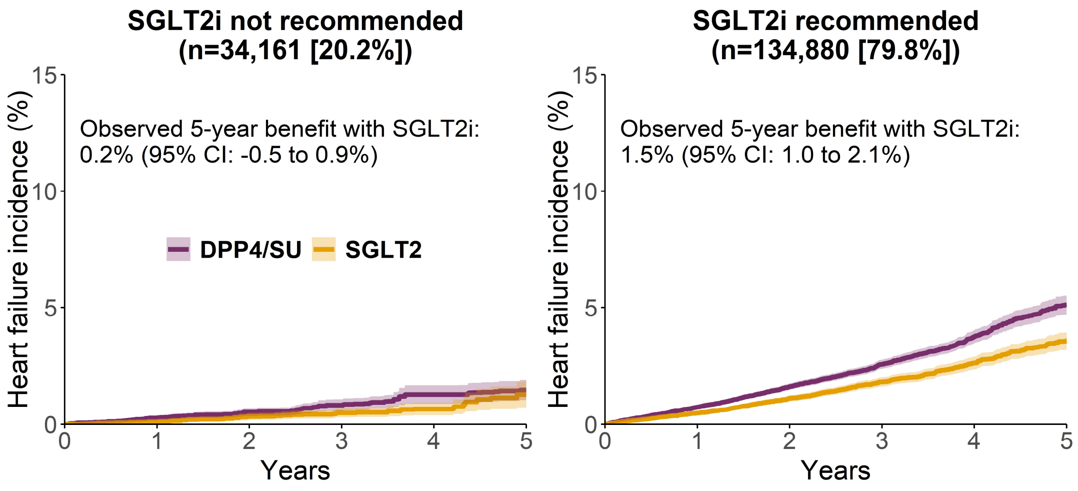


### b) SABRE model matched to NICE

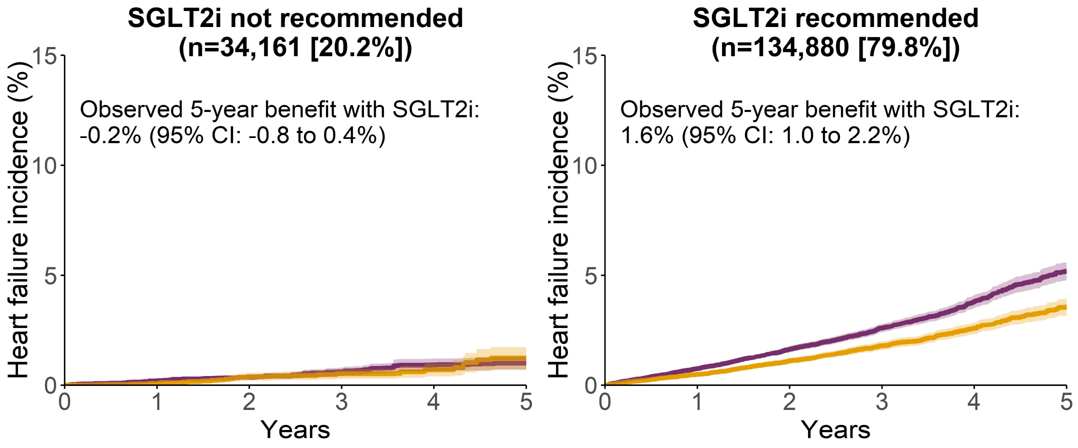

**c) SABRE model matched to ADA/EASD**

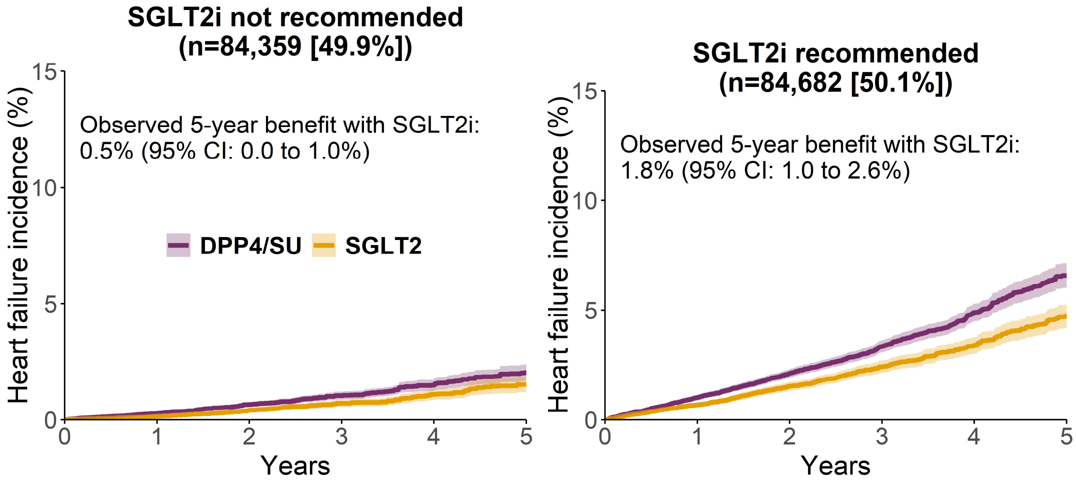


**d) QRISK2 model matched to ADA/EASD**

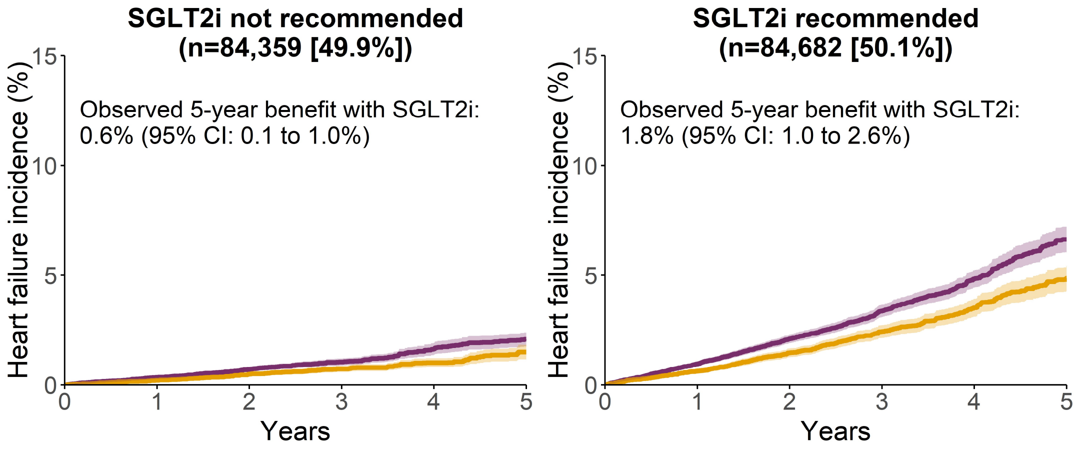


**e) QRISK2 restricted strategy**

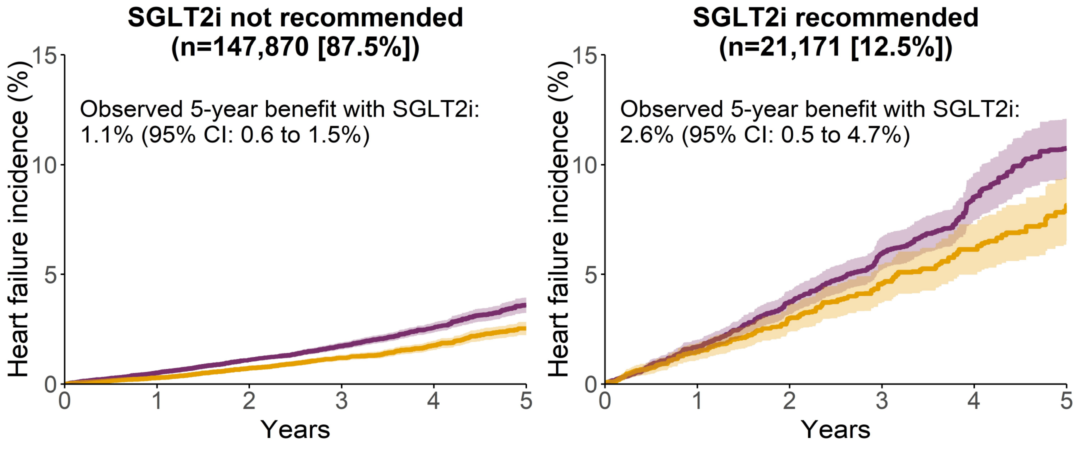


DPP4i=DPP4-inhibitor, SABRE= SGLT2i absolute benefit response model, SGLT2i=SGLT2i-inhibitor, SU=sulfonylurea.
